# The Genetic Landscape of Pediatric Postural Orthostatic Tachycardia Syndrome

**DOI:** 10.1101/2024.05.03.24306814

**Authors:** Huiqi Qu, Jingchun Qu, Xiao Chang, Nolan Williams, Frank Mentch, James Snyder, Maria Lemma, Kenny Nguyen, Meckenzie Behr, Michael March, John Connolly, Joseph Glessner, Jeffrey R Boris, Hakon Hakonarson

**Affiliations:** The Center for Applied Genomics, Children’s Hospital of Philadelphia, Philadelphia, Pennsylvania, 19104, USA; Department of Pediatrics, The Perelman School of Medicine, University of Pennsylvania, Philadelphia, Pennsylvania, 19104, USA; Division of Human Genetics, Children’s Hospital of Philadelphia, Philadelphia, Pennsylvania, 19104, USA; Jeffrey R. Boris, MD LLC, Moylan, PA 19065, USA; Division of Pulmonary Medicine Children’s Hospital of Philadelphia, Philadelphia, Pennsylvania, 19104, USA; Faculty of Medicine, University of Iceland, 101 Reykjavik, Iceland

**Keywords:** burden analysis, estrogen response, gene-based association, pathogenic variant, dysautonomia

## Abstract

**Background:** Postural orthostatic tachycardia syndrome (POTS) is a complex disorder with serious health consequences, while its etiology remains largely elusive.

**Objective:** To investigate the genetic landscape of POTS using genomic approaches in a unique pediatric cohort.

**Methods:** We conducted a combined genome wide genotyping and whole exome sequencing (WES) study to systemically examine the molecular mechanisms of POTS pathogenesis. The patients for were genotyped as two independent cohorts, a family cohort of 100 complete families and a case control cohort of 207 unrelated European cases and 4,063 ethnicity-matched controls. The WES component consisted of a subset of the genotyped subjects, including 87 unrelated European cases and 2,719 unrelated European controls.

**Results:** Due to the heterogeneous phenotype of POTS, unlike traditional phenotypes for association study, it is unlikely that any loci will reach genome-wide significance. Instead, we conducted an over-representation analysis (ORA) by considering all genes that showed nominal significance. The ORA identified gene sets linked to Cell-Cell Junction, Early Estrogen Response, and Substance-Related Disorders, with statistical significance. Moreover, WES revealed 55 genes with genome-wide significance through rare variant burden analysis, harboring 92 variants classified as pathogenic or likely pathogenic by ClinVar.

**Conclusions and Relevance:** This study showcases the complex interplay between common and rare genetic variants in POTS development, marking an pioneering step forward in deciphering its complex etiologies. The insights gained from this research enriches our understanding of POTS, offering new avenues for precise treatment strategies and highlighting the need for continued research in this area.

## Introduction

Postural orthostatic tachycardia syndrome (POTS) is a dysautonomia condition characterized by persistent excessive upright tachycardia upon assuming an upright position, without concurrent orthostatic hypotension^1–4^. Chronic orthostatic intolerance caused by POTS leads to severe functional impairment and psychological distress to the patients, seriously affecting patients’ lives. Patients with POTS often experience a wide array of symptoms, including severe lightheadedness, palpitations, cognitive impairment, debilitating fatigue, disruptions in sleep patterns, varying levels of pain, recurrent headaches, and gastrointestinal disturbances. POTS was only formally recognized as a distinct medical condition in 1993^5^. POTS affects about 0.2% to 1.0% of the US population, and is more frequently seen in women^1,2,6^. The connection with sex remains poorly understood. Both children and adults can be affected, while a majority of patients were diagnosed prior to reaching menopause^7,8^. The tachycardia in POTS can be due to any of the many factors affecting venous return and cardiac stroke volume, e.g. inability to maintain peripheral vascular tone, low blood volume, or increased pooling in the splanchnic circulation and extremities^9^. While anxiety is common, it is not considered as a significant causal factor. Despite extensive research, the complex pathophysiology of POTS remains only partially understood.

In light of the elusive nature of POTS’ etiology, our study employed an integrative OMICS approach to explore the molecular underpinnings of its pathogenesis in a unique pediatric cohort. Previous research has suggested a genetic link to POTS, such as the identification of the A457P mutation in the *SLC6A2* gene (encoding norepinephrine transporter) causing POTS^10^. In our study, genome-wide genotyping of common variants and exome sequencing for rare variants were applied. Unlike traditional genome-wide association study (GWAS) phenotypes, it is unlikely that any loci will reach genome-wide significance due to the heterogeneous phenotype of POTS. Increasing the sample size may not be effective as it also complicates the heterogeneity. Instead, focusing on common variants, we aimed to identify gene sets tagged by common single nucleotide polymorphisms (SNP) that are over-represented in POTS patients.

Through the exome sequencing, we aimed at identifying rare coding genetic variants in the candidate genes uncovered that may contribute to the disorder’s heterogeneous etiology. By centering on pediatric POTS patients, our study offers a unique perspective on the genetic landscape of this condition, potentially revealing insights into its complex mechanisms.

## Methods

### 1. GWAS

#### 1.1 Subjects

Participants were enrolled through the POTS Program at the Children’s Hospital of Philadelphia (CHOP). POTS patients aged 18 years or younger at the time of diagnosis were eligible for this study. We invited patients and their families to join this study through letters or emails. Those with DNA samples available from both parents were included in the family cohort. Unrelated patients lacking parental DNA samples were included in the case-control cohort. We obtained informed consent from all subjects or, if subjects are under 18, from a parent and/or legal guardian with assent from the child if 7 years or older. The CHOP Institutional Review Board (IRB) approved this study.

#### 1.2 Genotyping

The genotyping was done using the Illumina Infinium Global Screening Array (Illumina, San Diego, CA) with >700,000 SNPs genotyped. Altogether, 93.5% SNPs had a calling rate>99%, and the average calling rate of each DNA sample was 98.2%. Genome-wide imputation was done with the TOPMed Imputation Server (https://imputation.biodatacatalyst.nhlbi.nih.gov/<u>#</u>!) using the TOPMed (Version R2 on GRC38) Reference Panel. Altogether, 19,537,894 autosomal single nucleotide variants (SNV) with quality R^2^ ≥ 0.3 were included in this study.

#### 1.3. Genotyping data analysis

In this study, the family cohort was tested by transmission disequilibrium test (TDT), which is immune to spurious associations from population stratification^11^. Kinship between family members in the family cohort was validated by identity by descent (IBD) analysis based on the auto-chromosomal genotyping data. Loci with Mendelian errors > 3 were removed from association test. In addition, previous study has emphasized that replicated sequences in autosomes and sex chromosomes cause sex-related bias on the genotyping of autosomal SNPs^12^. As an additional quality filter, we tested sex effect by comparing mothers and fathers in the family cohorts. All SNPs with sex effect P<0.05 were removed. For the case-control cohort, unrelated cases were identified of European ancestry (EA) by principal component analysis (PCA) with genome-wide SNP markers, and were confirmed of non-relationship by identical-by-descent (IBD) analysis. European controls were selected by matching ethnicity based on the PCA analysis. Correction for population stratification was done by logistic regression using the first ten principal components as covariates^13^. The IBD analysis, TDT test, and case-control association test, were done using the PLINK software v1.9 (http://pngu.mgh.harvard.edu/purcell/plink/)^14^. All SNPs with Hardy-Weinberg Equilibrium (HWE) P<0.01 in European controls were removed from further analysis. Gene-based association test was done by the Versatile Gene-based Association Study - 2 version 2 (VEGAS2v02) software^15,16^.

### 2. WES

#### 2.1 Subjects

Based on the genotyping data, we identified 87 unrelated European cases (61 females and 26 males) with non-relationship validated by the IBD analysis, and European ancestry validated by the PCA analysis. The cases were compared with the Non-Finish European (NFE) population in the Exome Aggregation Consortium (ExAC) database^17^, using the Test Rare vAriants with Public Data (TRAPD) software^18^. Considering the potential inflation with the public database controls, the cases were further compared with 2,719 unrelated European non-POTS controls that have been sequenced by WES at the Center for Applied Genomics (CAG) of the Children’s Hospital of Philadelphia (CHOP).

#### 2.2 Library Preparation and Sequencing

Paired-end sequencing was performed on the Illumina NovaSeq 6000 platform (Illumina, San Diego, CA), using an S4 flowcell with run parameters of 101 x 10 x 10 x101 [Read 1 x Index 1 (i7) x Index 2 (i5) x Read 2]. Demultiplexing, alignment, and variant calling processes were performed on the Illumina DRAGEN Bio-IT Platform (version 3.3.7) using the 1000 Genomes Project Reference Human Genome Sequence (hs37d5). Alignment metrics were calculated using the Picard (version 2.18.27) CollectHsMetrics tool.

#### 2.3 Burden analysis of variants of interest (VOI) and pathogenic (P) or likely pathogenic (LP) Variants

The genetic variants which have minor allele frequency (MAF) greater than 0.001 in the NFE population based on the ExAC database^17^ have been excluded. Functional candidate VOIs were selected by the prediction results with at least 1 of a number of genetic variant prediction softwares, i.e. SIFT_pred="D" or Polyphen2_HDIV_pred="D" or Polyphen2_HDIV_pred="P" or Polyphen2_HVAR_pred="D" or Polyphen2_HVAR_pred="P" or LRT_pred="D" or MutationTaster_pred="A" or MutationTaster_pred="D" or MutationAssessor_pred="H" or MutationAssessor_pred="M" or FATHMM_pred="D" or PROVEAN_pred="D" or MetaSVM_pred="D" or MetaLR_pred="D", based on the annotation with the ANNOVAR software ^19^. The mutation burden in cases and controls were counted with the TRAPD software^18^. We have optimized the TRAPD algorithm with normalized genome coverage to capture causal variants with effects in the same directions.^20^ Gene-wide burden test of the candidate variants in the cases was done by one tailed Fisher exact test, compared with the ExAC NFE controls by dominant inheritance model. Multiple comparisons were corrected by Bonferroni correction. Assuming 21,306 protein-coding genes in human genome^21^, the genome-wide significance of the burden test was defined as α=0.05/21,306=2.347E-06. In further, deleterious variants were identified from the functional candidate variants according to the aggregated information by ClinVar annotation^22,23^, InterVar prediction^24^, and the Human Gene Mutation Database (HGMD) classification^25^.

## Results

### 1. Subjects

All POTS patients in this study were Caucasian with the age of diagnosis ranging from 12 years old to 21 years old, and median diagnosis age (Q1, Q3) of 15.6 (13.2, 17.8) years^7^. The patients were evaluated as two independent cohorts, a family cohort and a case control cohort. The family cohort included 114 POTS cases (including 28 males and 86 females) from 100 complete families. We included 62 unaffected siblings from these families in this study.

Significant comorbidities were detected in 18 unrelated patients from the family cohort (Table 1). The case control cohort included 207 unrelated cases (including 53 males and 154 females). Among the 207 cases, comorbidities were seen in 24 unrelated patients (Table 1). From the 207 cases, 194 unrelated cases (including 44 males and 150 females) based on genome-wide genotyping were compared with 4,063 European controls for genetic association test.

**Table 1.** Comorbidities with POTS.

| **Cohort** | **Comorbidities** | **Number of patients** |
| --- | --- | --- |
| **The family cohort (case n=114)** |  |  |
|  | Ehlers–Danlos Syndrome (EDS) | 11 |
|  | EDS + autoimmune alopecia | 1 |
|  | Mast cell activation syndrome (MCAS) | 1 |
|  | MCAS + Wolff-Parkinson-White syndrome + Leigh disease | 1 |
|  | Scoliosis+ Hashimoto's thyroiditis + benign premature atrial contractions | 1 |
|  | Crohn's disease | 1 |
|  | Post concussion | 1 |
|  | Benign Rolandic epilepsy | 1 |
| **The Case Control cohort (case n=207)** |  |  |
|  | EDS | 6 |
|  | EDS + eosinophilic esophagitis | 1 |
|  | EDS + Gilbert syndrome | 1 |
|  | EDS + IgA deficiency | 1 |
|  | EDS + MCAS | 1 |
|  | EDS + Chiari malformation + exercise-induced asthma + Asperger syndrome + gastroesophageal reflux + urticaria + and left duplicated ureter | 1 |
|  | Multiple sclerosis | 2 |
|  | MCAS | 1 |
|  | Crohn's disease | 1 |
|  | Alport's syndrome | 1 |
|  | Asperger syndrome + seizure disorder | 1 |
|  | Behçet's disease | 1 |
|  | Post concussion | 1 |
|  | Hodgkin lymphoma | 1 |
|  | Type 1 diabetes | 1 |
|  | Neuromuscular disorder | 1 |
|  | Congenital adrenal hyperplasia + von Willebrand's disease + Hashimoto's disease | 1 |
|  | UTI + VUR + asthma + vitamin D insufficiency + Lyme disease | 1 |

### 2. GWAS results

The inherently heterogeneous phenotype of POTS presented significant challenges. Achieving genome-wide significance for any particular genetic loci was improbable by increasing sample size. Instead, we conducted an over-representation analysis (ORA) by considering all genes that showed nominal significance. Our conclusions were only drawn based on ORA analysis with solid statistical evidence. In this study, 5,670 SNPs were identified of potential association signals with P<0.05 in both the family cohort and the case-control cohort, with effects in the same direction. The summary statistics are available in Supplementary Table 1. As shown, none of these loci showed genome-wide significance. Consequently, we performed gene-based association test at these loci. As shown in Supplementary Table 2, 716 genes showed association P<0.05 in both the family cohort and the case-control cohort, a number significantly higher (P=1.81E-128) than the 53 genes expected by chance (i.e., 21,306 × 5% × 5%), assuming there are 21,306 human coding genes.

Using the WebGestalt (WEB-based Gene SeT AnaLysis Toolkit) web tool^26^, over-representation analysis (ORA) of the 716 genes by the DisGeNET approach^27^, the Gene Ontology (GO) Cellular Component^28^, the GO molecular function^28^, the MSigDB Hallmark gene sets^29^, and the Human Phenotype Ontology (HPO)^30^, underscored several gene sets of statistical significance (FDR<0.1), with common genetic variants contributing to the susceptibility of POTS (Table 2).

**Table 2.**
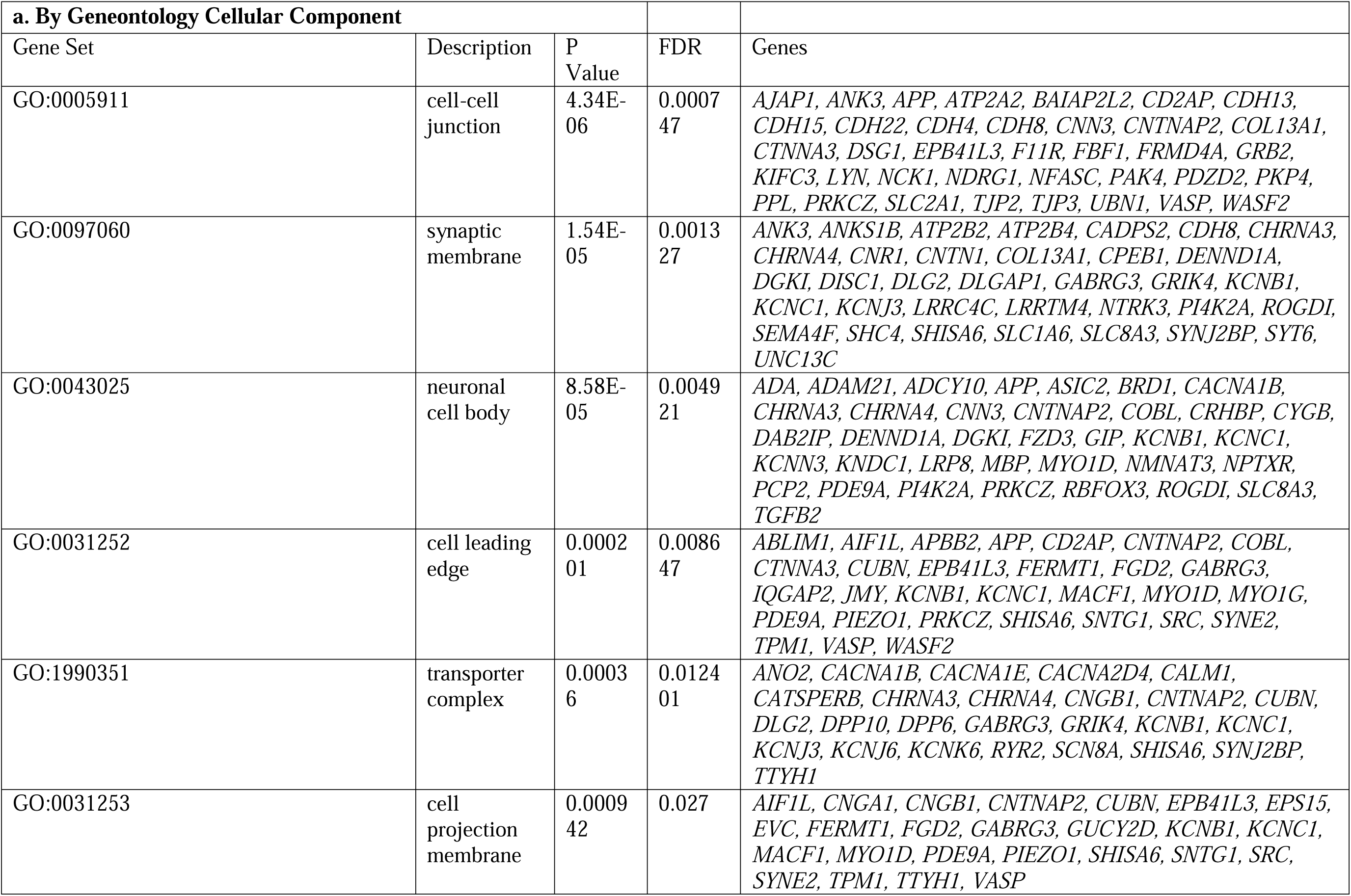
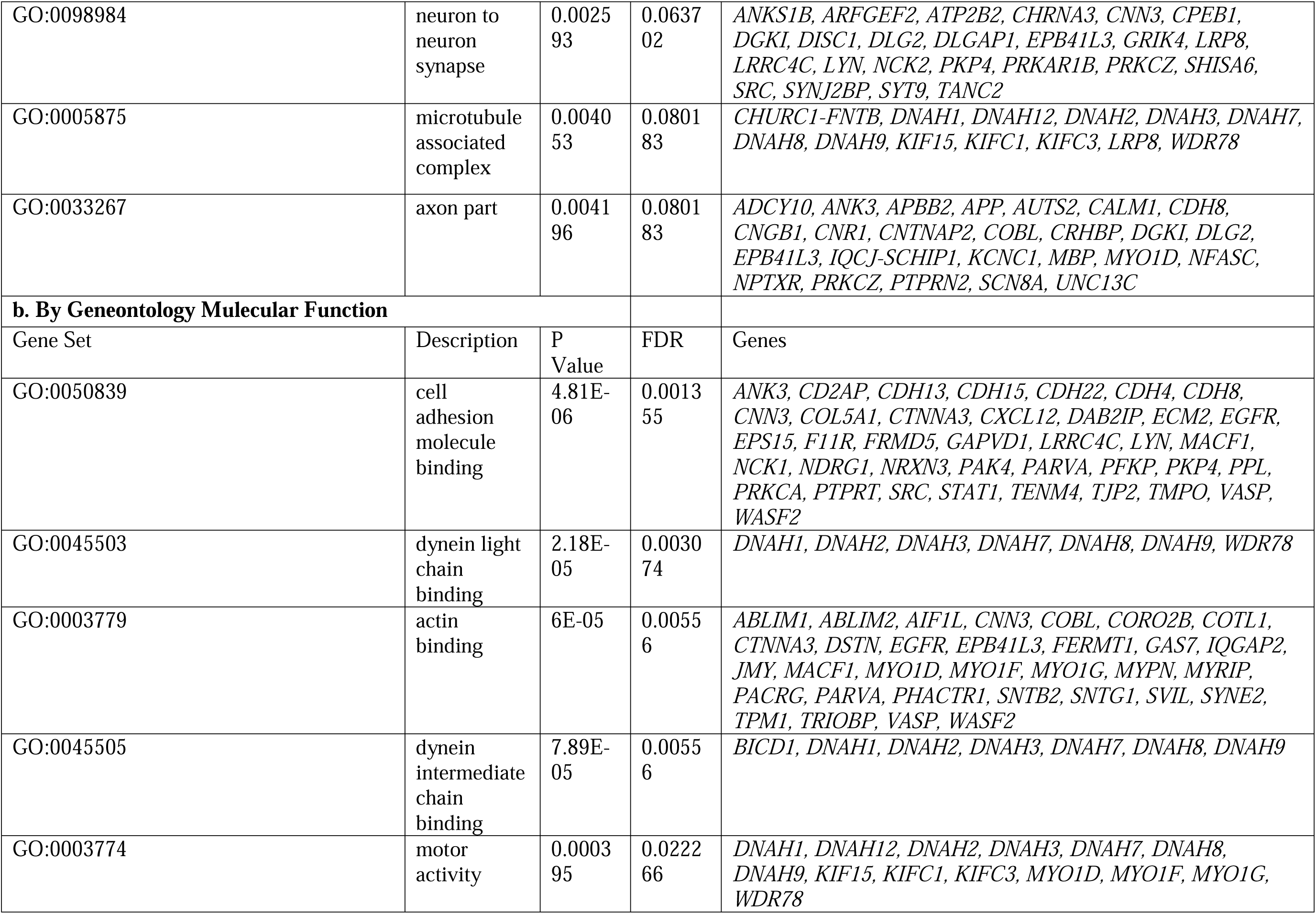
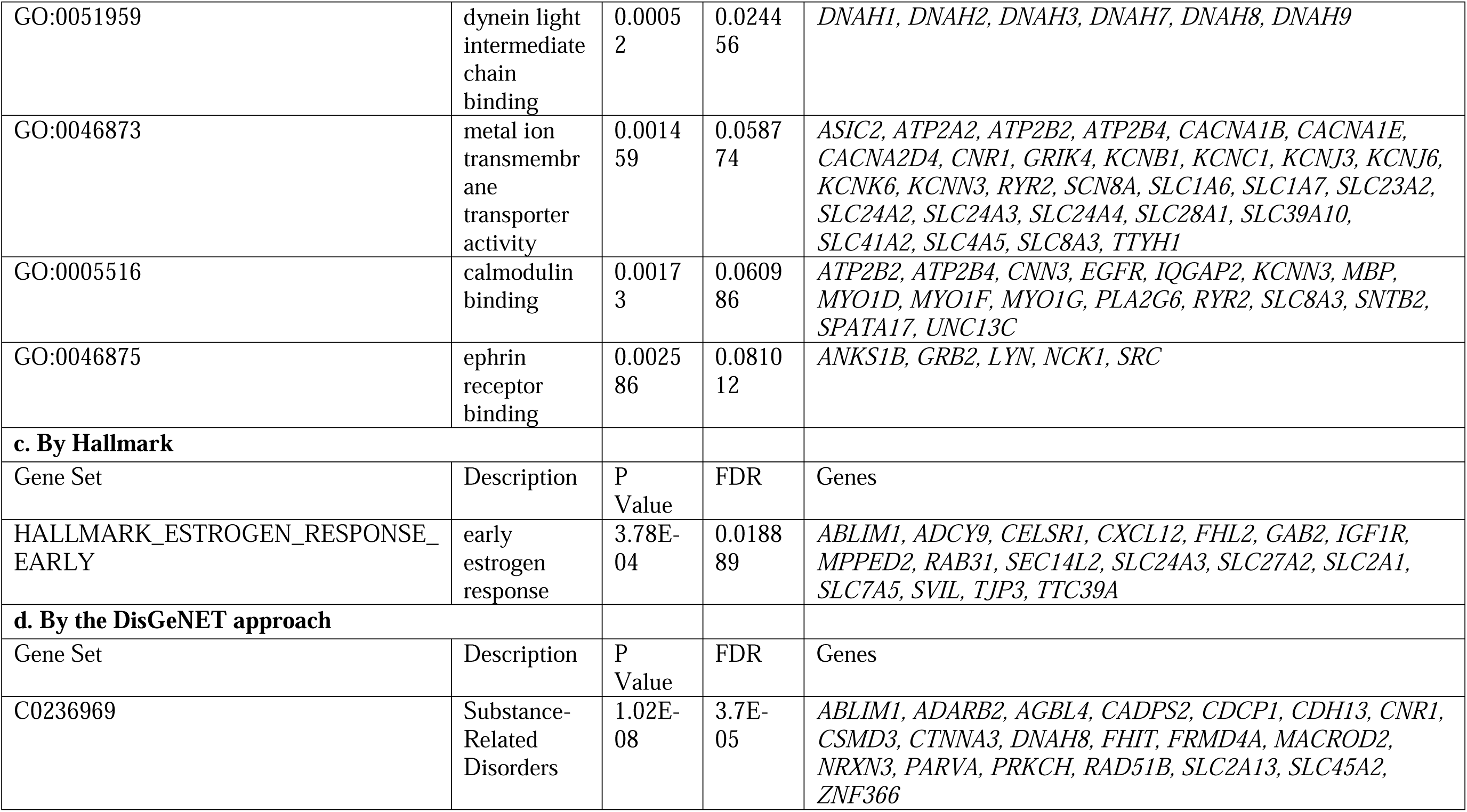
Over-representation analysis of the 716 genes showed nominal significance in both the family cohort and the case-control cohort.

### 3. Burden analysis of VOIs

By WES, 10,199 functional rare coding variants from 6,566 autosomal genes were called and annotated using the ANNOVAR software^19^. The gene burden analysis of these variants is shown in Supplementary Table 3. Considering the potential inflation with public database controls by TRAPD^18^, the cases were further compared with 2,719 unrelated European non-POTS controls that have been sequenced by WES at CAG. Significant genes were defined as genome-wide significant by comparing to both public database controls and the internal controls. As the results, 55 genes showed genome-wide significance (P<2.347E-06, Table 3). Among the 55 genes, 7 genes (*ABCA13, CELSR1, DAB2IP, DNAH1, DNAH2, DNAH3, SYNE2*) had nominal significance in the gene-based GWAS study [gene enrichment: 7/716 (gene-based GWAS) vs 55/21,306 (human coding genes), OR=3.81, P=3.52E-04], which suggests the association signals of these genes in GWAS may be explain by rare coding variants. ORA analysis of the 55 genes highlighted the roles of the genes in muscular function, emphasizing muscular dysfunction in the pathogenesis of POTS (Table 4).

**Table 3.** The 55 Genes showed genome-wide significance by burden analysis of rare coding variants.

| #GENE | CASE_COUNT_HET | CASE_COUNT_CH | CASE_COUNT_HOM | Control_COUNT_HET | Control_COUNT_CH | Control_COUNT_HOM | P_DOM |
| --- | --- | --- | --- | --- | --- | --- | --- |
| *ABCA13* | 10 | 2 | 0 | 26 | 7 | 0 | 6.40E-08 |
| *CELSR1* | 5 | 0 | 0 | 2 | 0 | 0 | 5.11E-07 |
| *DAB2IP* | 8 | 0 | 0 | 8 | 0 | 0 | 6.50E-09 |
| *DNAH1* | 9 | 0 | 0 | 20 | 0 | 0 | 1.05E-07 |
| *DNAH2* | 7 | 0 | 0 | 13 | 0 | 0 | 1.21E-06 |
| *DNAH3* | 9 | 1 | 0 | 25 | 4 | 1 | 6.37E-07 |
| *SYNE2* | 7 | 0 | 0 | 14 | 0 | 0 | 1.77E-06 |
| *ABCA7* | 7 | 1 | 0 | 8 | 6 | 0 | 1.14E-07 |
| *AGRN* | 6 | 0 | 0 | 5 | 0 | 0 | 3.05E-07 |
| *AHNAK* | 8 | 2 | 0 | 21 | 0 | 0 | 1.56E-06 |
| *AP5Z1* | 5 | 0 | 0 | 3 | 0 | 0 | 1.33E-06 |
| *ARHGA*<br>*P22* | 4 | 0 | 0 | 0 | 0 | 0 | 8.64E-07 |
| *ARID1B* | 5 | 0 | 0 | 3 | 0 | 0 | 1.33E-06 |
| *BAHCC*<br>*1* | 9 | 0 | 0 | 5 | 0 | 0 | 3.08E-11 |
| *CACNA*<br>*1A* | 5 | 1 | 0 | 3 | 1 | 0 | 1.33E-06 |
| *CACNA*<br>*1D* | 6 | 0 | 0 | 8 | 0 | 0 | 1.84E-06 |
| *CFAP46* | 6 | 0 | 0 | 6 | 0 | 0 | 5.95E-07 |
| *CILP* | 4 | 0 | 0 | 0 | 0 | 0 | 8.64E-07 |
| *CMYA5* | 4 | 0 | 0 | 0 | 0 | 0 | 8.64E-07 |
| *COL12A*<br>*1* | 5 | 0 | 0 | 2 | 0 | 0 | 5.11E-07 |
| *COL27A*<br>*1* | 6 | 0 | 0 | 5 | 0 | 0 | 3.05E-07 |
| *COL7A1* | 8 | 0 | 0 | 9 | 0 | 0 | 1.20E-08 |
| *CSMD1* | 6 | 0 | 0 | 6 | 0 | 0 | 5.95E-07 |
| *DNAH10* | 7 | 0 | 0 | 5 | 0 | 0 | 1.51E-08 |
| *EPHB4* | 5 | 0 | 0 | 1 | 0 | 0 | 1.50E-07 |
| *FAT1* | 6 | 0 | 0 | 8 | 0 | 0 | 1.84E-06 |
| *FBXW5* | 6 | 2 | 0 | 2 | 1 | 0 | 1.99E-08 |
| *FIGNL1* | 6 | 0 | 0 | 7 | 0 | 0 | 1.08E-06 |
| *FLG* | 7 | 0 | 0 | 5 | 0 | 0 | 1.51E-08 |
| *HSPG2* | 11 | 1 | 0 | 25 | 0 | 0 | 4.29E-09 |
| *KMT2C* | 8 | 0 | 0 | 8 | 0 | 0 | 6.50E-09 |
| *KNTC1* | 5 | 0 | 0 | 2 | 0 | 0 | 5.11E-07 |
| *LAMA5* | 13 | 1 | 0 | 21 | 0 | 0 | 5.38E-12 |
| *LRP2* | 11 | 0 | 0 | 11 | 0 | 0 | 7.17E-12 |
| *MUC16* | 14 | 2 | 0 | 25 | 0 | 0 | 2.11E-12 |
| *MYH7B* | 5 | 0 | 0 | 2 | 0 | 0 | 5.11E-07 |
| *NEB* | 12 | 3 | 0 | 21 | 1 | 0 | 7.64E-11 |
| *NRAP* | 5 | 0 | 0 | 3 | 0 | 0 | 1.33E-06 |
| *NUP160* | 4 | 0 | 0 | 0 | 0 | 0 | 8.64E-07 |
| *OBSCN* | 11 | 1 | 0 | 14 | 0 | 0 | 4.20E-11 |
| *PABPC1L* | 4 | 0 | 0 | 0 | 0 | 0 | 8.64E-07 |
| *PKD1* | 7 | 1 | 0 | 4 | 0 | 0 | 6.46E-09 |
| *PKD1L2* | 8 | 1 | 0 | 19 | 10 | 2 | 1.56E-06 |
| *PKHD1L1* | 7 | 0 | 0 | 9 | 0 | 0 | 1.97E-07 |
| *PLEC* | 11 | 1 | 0 | 21 | 0 | 0 | 1.02E-09 |
| *PLXNA2* | 4 | 0 | 0 | 0 | 0 | 0 | 8.64E-07 |
| *RP1L1* | 6 | 0 | 0 | 8 | 1 | 0 | 1.84E-06 |
| *RYR1* | 8 | 0 | 0 | 5 | 0 | 0 | 7.02E-10 |
| *SACS* | 8 | 0 | 0 | 4 | 0 | 0 | 2.77E-10 |
| *SRRM2* | 5 | 0 | 0 | 2 | 0 | 0 | 5.11E-07 |
| *TG* | 6 | 0 | 0 | 5 | 0 | 0 | 3.05E-07 |
| *TTN* | 24 | 5 | 0 | 51 | 1 | 0 | 1.72E-19 |
| *USH2A* | 9 | 0 | 0 | 9 | 0 | 0 | 6.76E-10 |
| *XIRP2* | 6 | 0 | 0 | 7 | 0 | 0 | 1.08E-06 |
| *ZFHX3* | 6 | 0 | 0 | 6 | 0 | 0 | 5.95E-07 |

Abbreviations: HET, heterozygote; CH, compound heterozygote; HOM, homozygote; P\_dom, P value of dominant model; P\_rec, P value of recessive model.

**Table 4.** Over-representation analysis of the 55 genes burdened with VOIs.

| **a. By the DisGeNET approach** |  |  |  |  |
| --- | --- | --- | --- | --- |
| Gene Set | Description | P Value | FDR | Genes |
| C1864711 | Muscle biopsy shows dystrophic changes | 2.41E-05 | 0.045614 | *PLEC, RYR1, SYNE2, TTN* |
| C0026850 | Muscular Dystrophy | 3.92E-05 | 0.045614 | *PLEC, RYR1, SYNE2, TTN* |
| C0221629 | Proximal muscle weakness | 5.55E-05 | 0.045614 | *NEB, RYR1, SYNE2, TTN* |
| C1838869 | Proximal neurogenic muscle weakness | 5.55E-05 | 0.045614 | *NEB, RYR1, SYNE2, TTN* |
| C0746674 | Generalized muscle weakness | 0.000109 | 0.045614 | *NEB, PLEC, RYR1, TTN* |
| C0151576 | Elevated creatine kinase | 0.000113 | 0.045614 | *HSPG2, PLEC, RYR1, SYNE2, TTN* |
| C0241005 | Creatine phosphokinase serum increased | 0.000113 | 0.045614 | *HSPG2, PLEC, RYR1, SYNE2, TTN* |
| C0376175 | Bell Palsy | 0.000117 | 0.045614 | *COL12A1, NEB, RYR1, TTN* |
| C1858719 | Facial muscle weakness of muscles innervated by CN VII | 0.000117 | 0.045614 | *COL12A1, NEB, RYR1, TTN* |
| C0427055 | Facial Paresis | 0.000125 | 0.045614 | *COL12A1, NEB, RYR1, TTN* |
| **b. By Geneontology Cellular Component** |  |  |  |  |
| Gene Set | Description | P Value | FDR | Genes |
| GO:0043292 | contractile fiber | 3.00E-10 | 5.16E-08 | *AHNAK, CACNA1D, CMYA5, MYH7B, NEB, NRAP, OBSCN, PLEC, RYR1, SYNE2, TTN, XIRP2* |
| GO:0042383 | sarcolemma | 3.78E-04 | 0.018834 | *AHNAK, CACNA1D, OBSCN, PLEC, RYR1* |
| GO:0031012 | extracellular matrix | 4.02E-04 | 0.018834 | *AGRN, CILP, COL12A1, COL27A1, COL7A1, FLG, HSPG2, LAMA5, USH2A* |
| GO:0016528 | sarcoplasm | 4.38E-04 | 0.018834 | *CMYA5, PLEC, RYR1, SYNE2* |
| GO:0030055 | cell-substrate junction | 5.55E-04 | 0.019084 | *AHNAK, ARHGAP22, FAT1, HSPG2, NRAP, PLEC, SYNE2, XIRP2* |
| **c. By Geneontology Molecule Function** |  |  |  |  |
| Gene Set | Description | P Value | FDR | Genes |
| GO:0005201 | extracellular matrix structural constituent | 9.23E-07 | 0.000197 | *AGRN, CILP, COL12A1, COL27A1, COL7A1, HSPG2, LAMA5* |
| GO:0045503 | dynein light chain binding | 1.40E-06 | 0.000197 | *DNAH1, DNAH10, DNAH2, DNAH3* |
| GO:0051959 | dynein light intermediate chain binding | 2.60E-06 | 0.000211 | *DNAH1, DNAH10, DNAH2, DNAH3* |
| GO:0045505 | dynein intermediate chain binding | 3.00E-06 | 0.000211 | *DNAH1, DNAH10, DNAH2, DNAH3* |
| GO:0042805 | actinin binding | 8.80E-06 | 0.000496 | *CACNA1D, NRAP, TTN, XIRP2* |
| GO:0008307 | structural constituent of muscle | 1.43E-05 | 0.000674 | *NEB, OBSCN, PLEC, TTN* |
| GO:0030506 | ankyrin binding | 4.03E-05 | 0.001624 | *CACNA1D, OBSCN, PLEC* |
| GO:0003774 | motor activity | 9E-05 | 0.003173 | *DNAH1, DNAH10, DNAH2, DNAH3, MYH7B* |
| GO:0003779 | actin binding | 0.000481 | 0.01508 | *MYH7B, NEB, NRAP, PLEC, SYNE2, TTN, XIRP2* |
| GO:0016887 | ATPase activity | 0.000627 | 0.017687 | *ABCA13, ABCA7, DNAH1, DNAH10, DNAH2, DNAH3, FIGNL1* |
| **d. By Hallmark** |  |  |  |  |
| Gene Set | Description | P Value | FDR | Genes |
| None |  |  |  |  |
| **e. By Human Phenotype Ontology** |  |  |  |  |
| Gene Set | Description | P Value | FDR | Genes |
| HP:0003306 | Spinal rigidity | 4.94E-07 | 0.002311 | *AGRN, COL12A1, HSPG2, NEB, SYNE2, TTN* |
| HP:0003458 | EMG: myopathic abnormalities | 3E-05 | 0.034381 | *AGRN, COL12A1, NEB, RYR1, SYNE2, TTN* |
| HP:0003457 | EMG abnormality | 3.84E-05 | 0.034381 | *AGRN, CACNA1D, COL12A1, HSPG2, NEB, RYR1, SYNE2, TTN* |
| HP:0003701 | Proximal muscle weakness | 0.000042 | 0.034381 | *AGRN, COL12A1, NEB, PLEC, RYR1, SYNE2, TTN* |
| HP:0100285 | EMG: impaired neuromuscular transmission | 4.27E-05 | 0.034381 | *AGRN, CACNA1D, RYR1, TTN* |
| HP:0003324 | Generalized muscle weakness | 4.41E-05 | 0.034381 | *AGRN, COL12A1, NEB, PLEC, RYR1, TTN* |
| HP:0003236 | Elevated serum creatine phosphokinase | 6.24E-05 | 0.03722 | *AP5Z1, COL12A1, HSPG2, NEB, PLEC, RYR1, SYNE2, TTN* |
| HP:0003690 | Limb muscle weakness | 6.48E-05 | 0.03722 | *AGRN, AP5Z1, NEB, RYR1, SACS, SYNE2, TTN* |
| HP:0040081 | Abnormal levels of creatine kinase in blood | 7.72E-05 | 0.03722 | *AP5Z1, COL12A1, HSPG2, NEB, PLEC, RYR1, SYNE2, TTN* |
| HP:0011021 | Abnormality of circulating enzyme level | 7.96E-05 | 0.03722 | *AP5Z1, COL12A1, HSPG2, NEB, PLEC, RYR1, SYNE2, TTN* |

### 4. Pathogenic/Likely Pathogenic (P/LP) variants

Our study identified 107 deleterious P/LP variants, of which 99 deleterious variants were supported by a minimum of two databases, including ClinVar (clinvar_20231230), InterVar, or HGMD_Pro_2023.3. When we concentrated on variants classified as P/LP variants by ClinVar classification, 92 P/LP variants were highlighted, including 3 variants that were previously reported of dominant genetic effects (Supplementary Table 4). Among the 87 WES patients, 53 (60.9%) have at least one P/LP variant (Supplementary Figure 1). The 92 P/LP variants are from 87 genes. Five genes, i.e., *GAA*, *GALT*, *GYS2*, *PAH*, and *USH2A*, each have two P/LP variants from two different individuals. The gene with P/LP variant, *OTOG*, has also been identified of nominal significance in the gene-based GWAS study. ORA analysis of the 87 genes highlighted a number of gene sets with statistical significance (FDR<0.1), suggesting the possibility of several underexplored gene pathways and networks in the pathogenesis of POTS (Table 5).

**Table 5.**
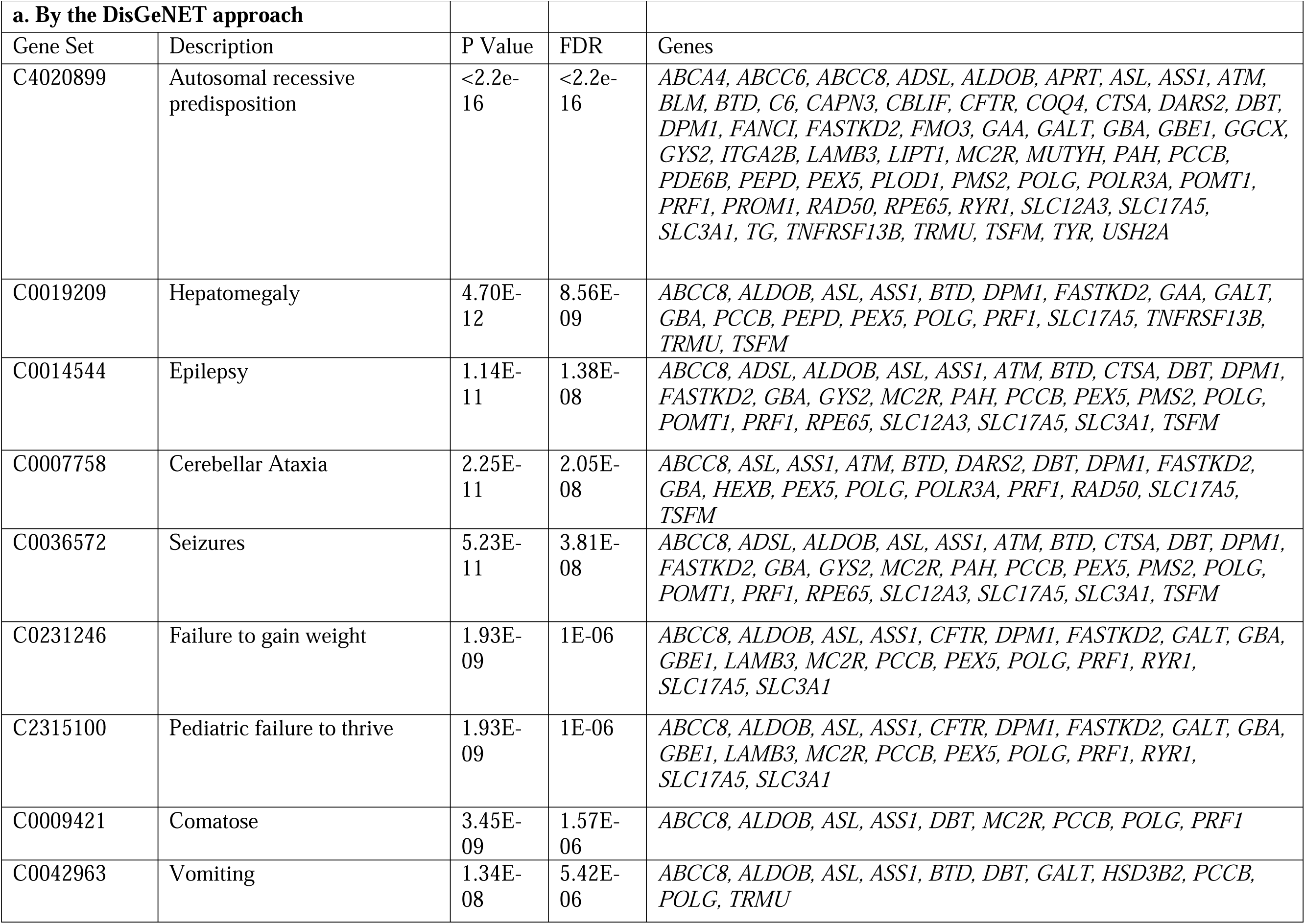
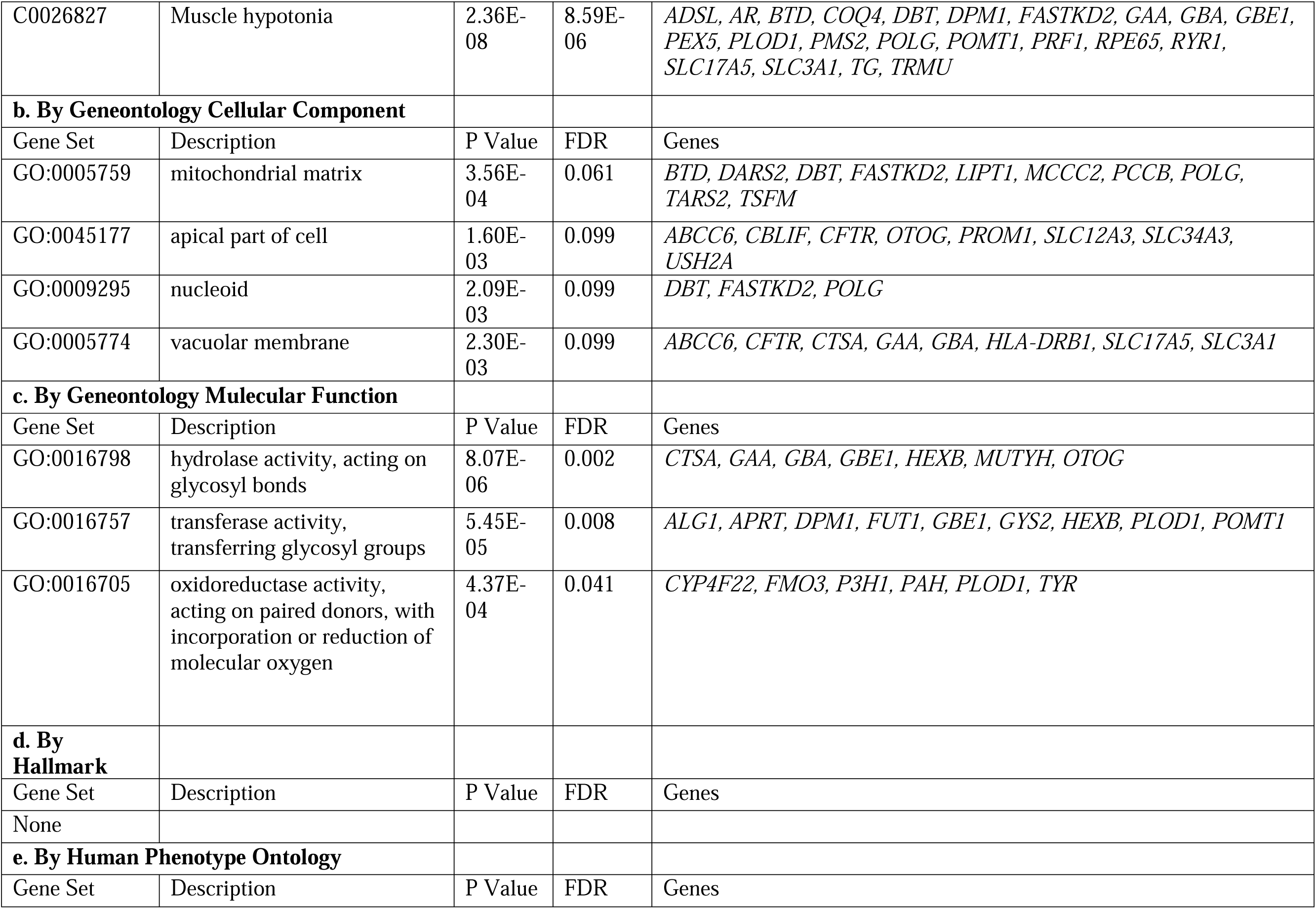
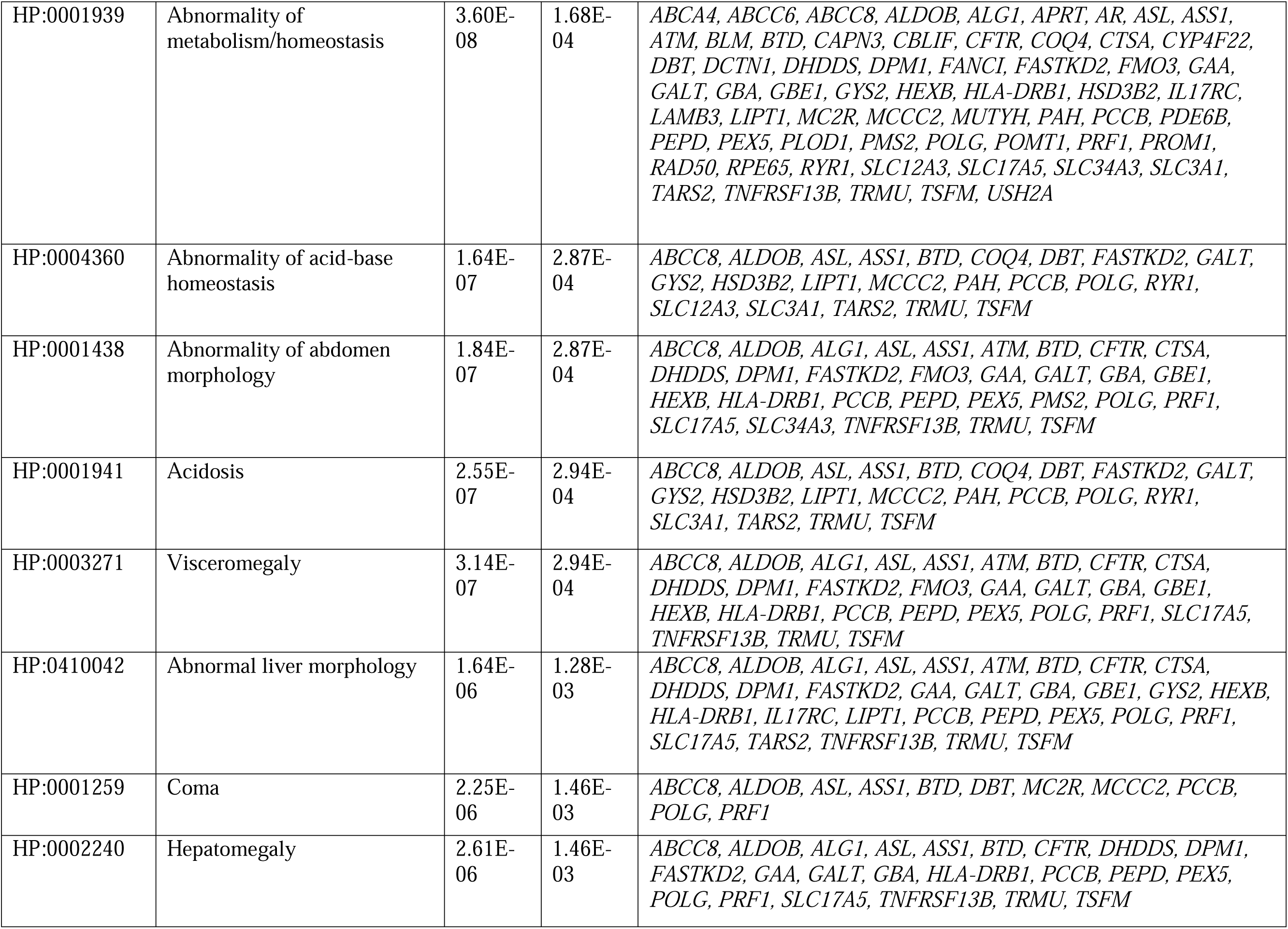
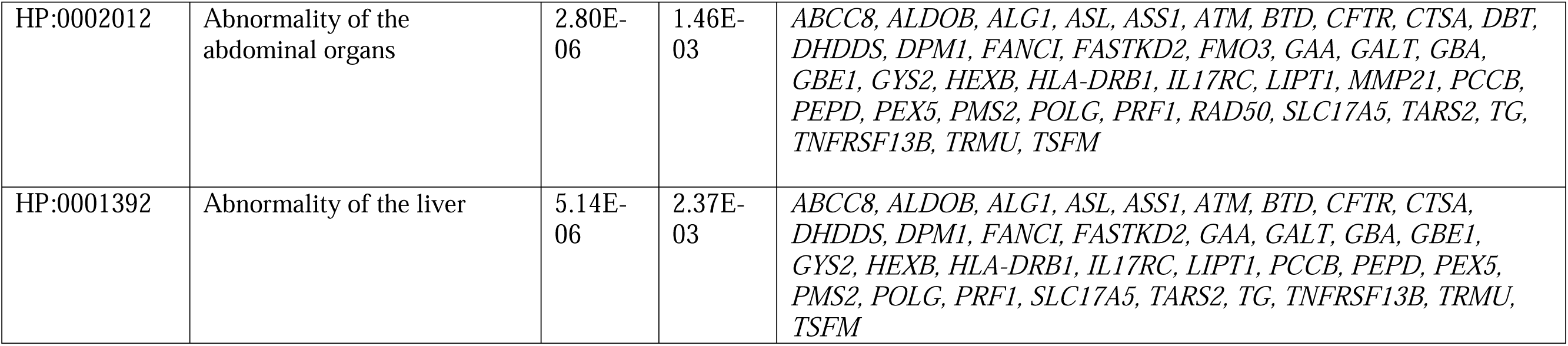
Over-representation analysis of the 87 genes with P/LP variants.

## Discussion

### 1. Common genetic variants and POTS susceptibility

This study presents a unique approach to a systemic evaluation of the etiology and molecular mechanisms of POTS. The application of GWAS to POTS has encountered challenges, primarily due to the disorder’s extensive phenotypic heterogeneity. This heterogeneity poses a significant obstacle for GWAS, which typically depends on a well-defined, uniform phenotype to effectively identify common genetic variants linked to a specific condition. A major challenge in GWAS for POTS is accurately characterizing its diverse phenotypes. The clinical complexity of POTS makes it difficult to distinguish between potential subtypes and to define a consistent phenotype that truly represents the disorder. Given the substantial phenotypic diversity of POTS, the GWAS approach was unable to identify any loci of genome-wide significance. Nevertheless, genes that showed nominal significance in gene-based association tests exhibited a highly significant enrichment in several gene sets important to POTS physiobiology. This finding underscores the role of common genetic variants in influencing POTS susceptibility and provides insights into its pathophysiology (Table 2).

#### 1.1 GO Cellular Component Cell-cell junction (GO:0005911) and synaptic membrane (GO:0097060)

These gene sets are integral to neuronal communication, which is crucial for the proper functioning of the ANS. Genes associated with cell-cell junctions play a role in maintaining the structural and functional integrity of synapses^31^, the points of communication between neurons. Synaptic membrane genes are involved in neurotransmitter release and reuptake^32^, which are critical for signal transmission in the ANS. Common genetic variations in genes associated with these processes can influence autonomic responses, a hallmark of POTS.

Neuronal cell body (GO:0043025) and axon part (GO:0033267): Genes associated with the neuronal cell body and axon are crucial for the health and function of neurons. Axonal genes play a role in the transmission of electrical signals along the nerve fiber^33^. Changed function in these cellular components by genetic variants can lead to impaired transmission of autonomic signals, contributing to the risk of orthostatic intolerance and tachycardia in POTS. Additionally, there is increasing evidence to suggest that a significant number of POTS patients experience small fiber neuropathy (SFN), an autoimmune disorder that specifically targets and damages the small fibers responsible for conducting autonomic and pain signals^34,35^. This further underscores the importance of understanding the genetic and cellular mechanisms underlying neuronal function and integrity.

Transporter complex (GO:1990351): This gene set is involved in the transport of various molecules across cellular membranes, including neurotransmitters^36^. In the context of POTS, the regulation of neurotransmitters like norepinephrine is particularly relevant. Dysregulation in neurotransmitter transport can lead to imbalances in sympathetic nervous system activity, a critical aspect of POTS pathophysiology^37^.

#### 1.2 GO Molecular Function Cell adhesion molecule binding (GO:0050839)

Genes involved in cell adhesion molecule binding play a crucial role in the interaction and adhesion of cells to their surrounding extracellular matrix and to other cells^38^. This is particularly important in the cardiovascular system, where endothelial cell integrity is essential for maintaining vascular function. In POTS, the dysregulation of this function could lead to compromised blood vessel reactivity and integrity, influencing blood flow dynamics.

Actin binding (GO:0003779): Actin is a fundamental component of the cellular cytoskeleton and is critical in various cellular processes, including maintenance of cell shape, cell movement, and muscle contraction^39^. Actin-binding genes are essential for the proper functioning of muscle cells, including cardiac^40^ and smooth muscle cells that line blood vessels^41^. In POTS, abnormalities in actin binding could impact cardiac muscle function and vascular tone regulation, both of which are vital for maintaining stable blood pressure and heart rate.

Motor activity (GO:0003774): This gene set is associated with the generation of force and movement within cells, a function that is crucial in muscle cells, including the heart^42^. In the context of POTS, motor activity genes could influence how heart and vascular muscles respond to autonomic signals, especially in adjusting heart rate and vascular tone in response to orthostatic stress.

#### 1.3 Early Estrogen Response (HALLMARK_ESTROGEN_RESPONSE_EARLY)

POTS is observed to be more common in women, with a ratio of as much as 5 females to 1 male^43^. However, the link with sex is not well comprehended. There is a recognized association between female hormones, notably estrogen, and changes in blood volume and vascular function^44^. This gene set comprises genes that are responsive to estrogen in the early phase of its action^45^. These early estrogen response genes could potentially play a role in POTS, given the higher prevalence of the condition in women. The potential effects include: (1) Autonomic regulation and cardiovascular effects: Estrogen is known to influence autonomic regulation and cardiovascular function^46^, which are both key aspects in the pathophysiology of POTS. (2) Extended Thoracic Hypovolemia: Estrogen can affect fluid retention and blood vessel constriction^47,48^, potentially influencing the degree of hypovolemia and the strain on the autonomic nervous system. (3) Autoimmune Responses: Estrogen can modulate immune responses^49^, which might intersect with autoimmune processes targeting the autonomic nervous system in POTS. Furthermore, females have a higher prevalence of autoimmune disorders compared to males^50^. (4) Inflammatory Mechanisms: Estrogen has both pro-inflammatory and anti-inflammatory effects, depending on the context^51^. The early estrogen response genes might play a role in the inflammatory underpinnings of POTS. (5) Autonomic Neuropathies and Sympathetic Denervation: Estrogen influences nerve function and repair^52^. Its early response genes could be involved in the development or compensation of autonomic neuropathies in POTS. (6) Impaired Norepinephrine Reuptake: Estrogen can modulate the expression and function of neurotransmitter transporters, possibly impacting norepinephrine reuptake mechanisms^53^. Clinically, we observed a case series of three transgender females transitioning to males whose POTS symptoms significantly improved after the addition of exogenous testosterone^54^. Additionally, both published^50^ and our unpublished data have observed that female POTS patients experience a worsening of symptoms around their menstrual periods.

#### 1.4 Substance-Related Disorders (DisGeNET C0236969)

This correlation carries two implications: firstly, POTS may share a common genetic susceptibility with substance-related disorders; secondly, this gene set might be linked to POTS due to the role of certain substances in modulating the autonomic nervous system and cardiovascular responses. Dysautonomia can be exacerbated or triggered by substance exposure. The underlying mechanisms may include: (1) The autonomic nervous system may be influenced by various medications commonly utilized in clinical practice^55^. For example, β-adrenergic receptors are activated by some bronchodilators for asthma management. Amphetamines, like those prescribed for attention deficit hyperactivity disorder, or consuming caffeine, can lead to an increase in the release of the sympathetic neurotransmitter norepinephrine. Tricyclic antidepressants can inhibit the reuptake of norepinephrine, thus increasing its availability in the synaptic cleft^56^. (2) Common substances can exert direct or indirect effects on the cardiovascular system, like caffeine, alcohol, nicotine, and antidepressants^57^. Calcium channel blockers may cause peripheral vasodilation and reduce venous return^58^, thus exacerbate the hypovolemic state often seen in POTS. β-blockers may influence myocardial contractility or heart rate, contributing to the dysregulation of cardiovascular function. (3) Substances can also alter the body’s response to stress, a factor that is often implicated in the exacerbation of POTS symptoms^59^. The dysregulation of stress hormones and the sympathetic nervous system can lead to increased heart rate and blood pressure variability.

These gene sets offer a window into the complex interplay of common genetic variants and their potential role in predisposing individuals to POTS. The exploration of GWAS gene sets in the context of POTS not only enhances our understanding of the genetic basis of the syndrome but also opens new pathways for personalized and preventive healthcare strategies.

### 2. Rare functional variants and POTS heterogeneity

Compared to the results of our GWAS study, our WES study emphasizes the importance of rare coding variants in the pathogenesis of POTS. Two complementary analyses were employed in this study: the burden analysis of rare variants and the identification of P/LP variants. The burden analysis entails assessing the cumulative impact of rare functional variants in the individuals with POTS compared to the control group. The primary focus is to determine whether there is a higher prevalence of functional rare variants in the POTS patients, as opposed to common variants identified in the association study. This analysis does not necessarily prioritize the predicted pathogenicity of each variant. Instead, it focuses on evaluating the overall burden of these functional rare variants in the genome, providing a comprehensive overview of the genetic landscape. Conversely, the analysis of P/LP rare variants involves identifying deleterious variants, particularly those classified by ClinVar. This can help establish a direct link between specific genetic changes and POTS, leading to a better understanding of the molecular mechanisms of the disease and potentially guiding targeted treatments.

#### 2.1 Insights gained by burden analysis of VOIs

Using 2,719 unrelated European controls, this study identified 55 genes associated with POTS with genome-wide significance by burden analysis of rare coding variants. The 55 genes identified in this study highlight both known and also unveil novel knowledge of POTS heterogeneity.

##### 2.1.1 Muscular dysfunction in POTS

The ORA analysis in this study emphasized the importance of possible muscular dysfunction in POTS, with genes involved in muscle function and muscular diseases enriched with highly statistical significance (Table 4a,e). Altogether, 32 out of the 55 genes are related to muscular dysfunction. The affected muscular function may not be limited to myocardium and vascular smooth muscle. For instance, the calf muscle pump generates pressure gradient between the thigh and the lower leg veins, and is the major force for return of venous blood from the lower extremities to the heart^60^. Decreased calf muscle pump activity (HP:0003690 Limb muscle weakness) may thus contribute to the venous pooling in lower extremities in some POTS patients^61^. It’s worth noting that no muscle dysfunction has been observed in these POTS patients, suggesting that any potential involvement of muscular mechanisms may be subclinical in terms of skeletal muscle dysfunction.

Muscular function relies on coordinated activity between muscle fibers and the metabolic and regulatory machineries^62^. The structural components of muscle cells that may be affected by rare coding variants include (Table 4b): (1) Contractile fiber (GO:0043292), sarcolemma (GO:0042383), and sarcoplasm (GO:0016528). The genes with rare coding variants include AHNAK nucleoprotein (*AHNAK*), calcium voltage-gated channel subunit alpha1 D (*CACNA1D*), cardiomyopathy associated 5 (*CMYA5*), myosin heavy chain 7B (*MYH7B*), nebulin (*NEB*), nebulin related anchoring protein (*NRAP*), obscurin, cytoskeletal calmodulin and titin-interacting RhoGEF (*OBSCN*), plectin (*PLEC*), ryanodine receptor 1 (*RYR1*), spectrin repeat containing nuclear envelope protein 2 (*SYNE2*), titin (*TTN*), and xin actin binding repeat containing 2 (*XIRP2*). (2) Extracellular matrix (GO:0031012). The genes with rare coding variants are agrin (*AGRN*), cartilage intermediate layer protein (*CILP*), collagen type XII alpha 1 chain (*COL12A1*), collagen type XXVII alpha 1 chain (*COL27A1*), collagen type VII alpha 1 chain (*COL7A1*), filaggrin (*FLG*), heparan sulfate proteoglycan 2 (*HSPG2*), laminin subunit alpha 5 (*LAMA5*), and usherin (*USH2A*). (3) Cell-substrate junction (GO:0030055). The related genes with rare coding variants are *AHNAK*, Rho GTPase activating protein 22 (*ARHGAP22*), FAT atypical cadherin 1 (*FAT1*), heparan sulfate proteoglycan 2 (*HSPG2*), *NRAP, PLEC, SYNE2*, and *XIRP2*. The molecular functions of these genes are related to the dynein motor to generate force, cytoskeletal actinin /ankyrin/actin binding, and ATPase activity for providing energy (Table 4c).

##### 2.1.2 Microtubule dysfunction in POTS

Among the 32 genes that are related to muscular dysfunction, four dynein axonemal heavy chain (DNAH) genes *DNAH1*, *DNAH2*, *DNAH3*, *DNAH10*, and the *SYNE2* gene involve microtubule function. Axonemal dynein produces force to move other proteins and cell materials by microtubules within cilia^63^. Dysfunction in endothelial cilia contributes to aberrant fluid-sensing and results in vascular disorders, including hypertension^64^. In addition, an intact microtubule network is necessary for proper subcellular structure and function^65^. Aberrant growth of cardiomyocyte microtubules contribute to contractile dysfunction^66^. Targeting at microtubules may improve cardiomyocyte function in human heart failure^67^. *SYNE2* encodes nuclear envelope spectrin-repeat protein (Nesprin)-2, functioning as intracellular scaffolds and linkers to establish nuclear-cytoskeletal connections by binding cytoplasmic F-actin, in addition to its role as a microtubule scaffold^68^. Mutations of *SYNE2* may lead to structural and adaptive signaling defects in mechanically stressed tissues such as muscle, and cause Emery-Dreifuss muscular dystrophy (EDMD5)^69^. Besides the above genes, two additional genes, kinetochore associated 1 (*KNTC1*) and RP1 like 1 (*RP1L1*) also encode proteins of the microtubule complex (GO:0005874). Notably, there has been no observed contractile dysfunction in these POTS patients, implying that any potential engagement of microtubule mechanisms may manifest subclinically concerning contractile function.

##### 2.1.3 Genes reported of association with blood pressure

According to the GWAS catalog, 15 of the 55 genes have been reported of association with blood pressure regulation (https://www.ebi.ac.uk, accessed on Sep 5, 2021), including 7 genes related to muscular function (*ARHGAP22, CACNA1D, DNAH2, DNAH3, PLEC, SACS, TTN*) with 8 other genes contributing (*ARID1B, BAHCC1, CSMD1, LRP2, NUP160, PKD1, RP1L1, ZFHX3*). The genes involved in muscular function may be related to POTS by their roles involving myocardium or vascular smooth muscle function. For example, the 2 DNAH genes *DNAH2*^70,71^ and *DNAH3*^72^ are associated with systolic blood pressure, while *DNAH3* is also reported of association with diastolic blood pressure^72^. The association of *DNAH2* and *DNAH3* with blood pressure may be related to their roles in cardiomyocyte function^66^ (for systolic blood pressure), and microtubule function in vascular smooth muscle contraction^73^. However, clinically, no contractile dysfunction has been demonstrated in POTS so far, suggesting the need for further investigation into the underlying mechanisms.

The LDL receptor related protein 2 gene (*LRP2*) encodes the endocytic receptor megalin, which has regulatory effects on the renin-angiotensin system activity in the kidney^74^, in addition to its key roles in renal proximal tubular function^75^.

##### 2.1.4 Genes reported of association with heart rate

Among the 55 genes, 10 genes have been reported of association with heart rate, including 4 genes related to muscular function (*CACNA1D, COL12A1, PLEC, TTN*) and 6 other contributing genes (*CELSR1, CSMD1, DAB2IP, EPHB4, RP1L1, ZFHX3*) (https://www.ebi.ac.uk, accessed on Sep 5, 2021). Among the 10 genes, the genes *CACNA1D, CSMD1, PLEC, RP1L1, TTN,* and *ZFHX3*, are also associated with blood pressure.

*DAB2IP* associated with heart rate^76^ encodes a Ras GTPase-activating protein. In addition to its role as a tumor suppressor^77^, *DAB2IP* protein functions as a scaffold protein and modulates different signal cascades associated with cell proliferation, survival, and apoptosis.^78^ Through the DAB2IP-ASK1-JNK signaling pathway, DAB2IP plays important roles in the function and apoptosis of vascular endothelial cells^79^.

*CELSR1* encodes a member of the flamingo subfamily of the cadherin superfamily^80^, with important roles in neuronal morphogenesis^81^. Mutations of this gene has been reported of correlation with neural tube defects^82^. *CELSR1* was reported of association with heart rate in heart failure patients by a previous GWAS^83^. Concerning the potential roles of *CELSR1* in regulating heart rate and in POTS, vestibular hair cells of the inner ear convert mechanical stimuli into neural activity, thus to control balance, blood pressure and heart rate^84^. *CELSR1* coordinates the planar polarity organization of vestibular hair cells in inner ear development^85^.

Meanwhile, we have observed patients who still have vestibular dysfunction clinically, even without a history of head trauma or concussion.

##### 2.1.5 Cardiac insufficiency resulting from genetic mutations

In addition to the knowledge gained from the gene set enrichment analysis, 12 of the 32 genes related to muscular dysfunction (*NEB, PLEC, XIRP2, TTN, CACNA1D, CMYA5, FAT1, HSPG2, MYH7B, NRAP, OBSCN, SYNE2*) have also been reported of association with cardiomyopathy according to the HGMD professional dataset^25^ 2021.1 release. Four of the 12 genes (*NEB, PLEC, XIRP2, TTN*) and 9 other genes (*ARID1B, CACNA1A, CELSR1, KMT2C, LRP2, AHNAK, COL7A1, LAMA5, RYR1*) are also related to congenital heart disease. For instance, the *TTN* gene encodes the giant muscle filament titin of striated muscle. *TTN* is associated with familial hypertrophic cardiomyopathy^86^ and familial dilated cardiomyopathy^87^, as well as a specific form of cardiomyopathy characterized by arrhythmia, i.e. arrhythmogenic right ventricular cardiomyopathy (ARVC)^88^. *MYH7B* encodes the major contractile protein in heart and vascular smooth muscle and is directly involved in muscle contraction^89^. These findings highlight a subset of POTS patients with rare coding variants from genes related to inherited cardiomyopathy, congenital heart defects, or congenital channelopathy (e.g. *RYR1*^90^, *CACNA1D*^91^). The POTS symptoms in these patients may be attributed to cardiac insufficiency resulting from genetic mutations, without necessarily involving subclinical or inconspicuous structural or functional changes.

##### 2.1.6 Psychiatric and Neurodevelopmental Disorders in POTS

It’s not uncommon for patients with POTS to experience psychological issues like depression and anxiety^92^. There is a potential bidirectional relationship between POTS and psychological distress, whereas the exact role of psychiatric and psychological factors in the development of POTS remains a topic of ongoing research. From the 55 genes we identified, 4 have been linked to anxiety disorder, 8 to schizophrenia, and 4 to autism spectrum disorder (ASD) (the GWAS catalog https://www.ebi.ac.uk, accessed on Sep 5, 2021). As per HGMD, 14 genes are linked to schizophrenia, and notably, 43 out of the 55 genes are related to ASD (Supplementary Table 5). Autonomic dysfunction is common in ASD^93^. The findings of our study imply that individuals diagnosed with POTS may also have concurrent atypical psychiatric or neurodevelopmental disorders. Owens et al. have documented a correlation between dysautonomia and ASD^94^. Moreover, in clinical settings, we have observed a number of POTS patients with ASD.

#### 2.2 Insights gained by ClinVar P/LP variants

In our WES study, we identified 92 heterozygous P/LP variants in 87 different genes classified by ClinVar. Many of these genes are associated with autosomal recessive predisposition; therefore, patients do not typically manifest obvious genetic syndromes when these variants are present in a heterozygous state. Among these genes, the otogelin gene (*OTOG*) has also been identified in the gene-based GWAS study on common genetic variants. *OTOG* encodes a protein that is primarily associated with the acellular membranes of the inner ear and plays a crucial role in auditory and vestibular functions^95^. The LP variant NP_001278992.1:p.Gly2238Ser causes a rare genetic deafness with autosomal recessive inheritance (https://www.ncbi.nlm.nih.gov/clinvar/variation/930161/). While *OTOG* is primarily associated with the inner ear, there is some evidence to suggest that ANS dysfunction can be linked to inner ear disorders^96^. Disruptions in the vestibular system can lead to balance and coordination problems, which may indirectly affect ANS regulation in some individuals. Furthermore, several intriguing genes offer additional insights into the pathogenesis of POTS.

##### 2.2.1 P/LP variants with Dominant effects

Among the 92 heterozygous P/LP variants, 3 have been reported of dominant genetic effects, including *USP48* (ubiquitin specific peptidase 48)*/*NP_115612.4:p.Gly406Arg causing Deafness, autosomal dominant 85; *CAPN3* (calpain 3)/NP_000061.1:p.Arg490Trp causing Muscular dystrophy, limb-girdle, autosomal dominant 4; *POLG* (DNA polymerase gamma, catalytic subunit)/NP_002684.1:p.Trp748Ser causing Progressive external ophthalmoplegia with mitochondrial DNA deletions, autosomal dominant 1. The co-occurrence of these P/LP variants with POTS could be coincidental. However, *CAPN3* encodes a muscle-specific component of the calpain protease, which is a muscle-specific member of the calpain large subunit family, and exhibiting a specific binding affinity for the protein titin^97^. The variant causing muscular dystrophy can lead to muscle weakness and mobility issues, contributing to POTS by promoting deconditioning and muscle pump dysfunction. *POLG* encodes the catalytic subunit of mitochondrial DNA polymerase, a critical enzyme responsible for replicating mitochondrial DNA^98^. POLG plays a pivotal role in maintaining the integrity and proper functioning of mitochondrial DNA, which is essential for the production of energy within cells. Mitochondrial dysfunction can affect multiple physiological processes, including those related to the autonomic nervous system and cardiovascular regulation, thus may contribute to POTS^99^. Besides these P/LP variants, the myosin heavy chain 7 (*MYH7*, related to hypertrophic cardiomyopathy) variant NP_000248.2:p.Arg787Cys at exon21 is classified as DM by HGMD and Likely pathogenic by InterVar, but with Conflicting interpretations of pathogenicity by ClinVar. *MYH7* encodes the beta (or slow) heavy chain subunit of cardiac myosin. This specific heavy chain is primarily expressed in the normal human ventricle, as well as in skeletal muscle tissues rich in slow-twitch type I muscle fibers^100^. Its mutation can affect myocardial contractility.

##### 2.2.2 Insights gained from enriched gene sets with P/LP variants

ORA analysis of the 87 genes with P/LP variants identified several gene sets of statistical significance. Significant DisGeNET gene sets include Hepatomegaly (C0019209), Epilepsy (C0014544), Cerebellar Ataxia (C0007758), Seizures (C0036572), Failure to gain weight (C0231246), Pediatric failure to thrive (C2315100), Comatose (C0009421), Vomiting (C0042963), Muscle hypotonia (C0026827). Hepatomegaly may be related to splanchnic redistribution of blood, contributing to thoracic hypovolemia in POTS. Epilepsy and seizures often cause autonomic nervous system dysfunction^101^. Cerebellar ataxia, affecting balance and coordination, may contribute to orthostatic intolenrance in POTS. Moreover, the association of POTS with gene sets linked to clinical diagnoses such as coma might suggest that certain genetic mutations have a profound impact on neurological functions. Muscle hypotonia can contribute to POTS by promoting deconditioning and muscle pump dysfunction.

Gene sets of GO Cellular Component include mitochondrial matrix (GO:0005759), and apical part of cell (GO:0045177). Dysfunction in the mitochondrial matrix can lead to energy deficits, which are implicated in dysautonomia and may impact muscle function, including the heart and vascular system, thus contribute to POTS^99^. The apical part of a cell is important in cellular polarization and signaling^102^. In endothelial cells, dysfunction in the apical part could affect vascular tone and blood flow regulation.

Gene sets of GO Molecular Function include hydrolase activity, acting on glycosyl bonds (GO:0016798), transferase activity, transferring glycosyl groups (GO:0016757), oxidoreductase activity, acting on paired donors, with incorporation or reduction of molecular oxygen (GO:0016705). Hydrolases that act on glycosyl bonds are involved in the breakdown of carbohydrates and glycoproteins^103^. Impaired carbohydrate metabolism could affect energy availability, potentially influencing the energy-dependent processes of the autonomic nervous system. Glycoproteins play roles in cell signaling and immune responses^104^. Abnormalities in glycoprotein breakdown could contribute to dysregulated immune responses, potentially relevant in autoimmune etiologies of POTS. Glycosylation is important in cell signaling and immune function^105^. Aberrations here could contribute to autoimmune responses or dysregulation of the autonomic nervous system, both implicated in POTS. Oxidoreductase enzymes play a central role in oxidative phosphorylation and energy production in cells, and are closely related to mitochondrial function. These enzymes also play roles in oxidative stress, which has been implicated in various pathologies, including inflammation and autoimmunity.

## Conclusion and perspective

Leveraging our expertise in omics and the analysis of heterogeneous phenotypes, this study marks an important step forward in understanding the complex etiologies of POTS, a condition with significant phenotypic heterogeneity and elusive genetic underpinnings. With convincing statistical significance, we have illuminated the role of both common and rare genetic variants in POTS development. We have identified several gene sets through GWAS, notably linked to cell-cell junctions, synaptic membranes, transporter complexes, and early estrogen responses. Our WES analysis brings into focus specific genes and molecular mechanisms including muscular and microtubule dysfunction, autonomic nervous system regulation, and mitochondrial activity. This enhanced genetic understanding opens new avenues for developing personalized treatment strategies, tailored to the unique genetic makeup of individual POTS patients. Meanwhile, the study’s findings regarding the relationship between POTS-related genes and psychiatric and neurodevelopmental disorders underscore the importance of addressing psychological aspects in the management of POTS.

The burden analysis of VOIs in the WES study identified 55 genes with statistical significance and 87 genes with P/LP variants (including 3 genes with dominant genetic variants). Due to the limitations imposed by the sample size and phenotypic heterogeneity, the GWAS study achieved statistical significance for several gene sets rather than for individual genes.

Nonetheless, common variants from several plausible candidate genes might exert regulatory effects as modifiers in the pathophysiology of POTS and merit further investigation. For instance, common variants in the glycoprotein alpha-galactosyltransferase 1 gene (*GGTA1*), the 3-oxoacid CoA-transferase 2 gene (*OXCT2*), and the 3-oxoacid CoA-transferase 2 pseudogene 1 gene (*OXCT2P1*), have shown nominal statistical significance in association with POTS. These variants are linked to gene expression in the heart atrial appendage, as per the Genotype-Tissue Expression (GTEx) project data (https://www.gtexportal.org/)^106^ (Supplementary Table 6), implying a direct role in heart rate regulation^107^, a key aspect of POTS pathogenesis. The genetic insights not only enhance our knowledge of POTS pathogenesis but also hold promise for developing more effective, individualized treatment strategies, ultimately improving patient outcomes in this challenging and multifaceted condition.

## Declarations

### Ethics approval and consent to participate

Informed consent was obtained from all subjects or, if subjects are under 18, from a parent and/or legal guardian with assent from the child if 7 years or older. The Institutional Review Board (IRB) of CHOP approved this study.

### Consent for publication

Not applicable.

### Availability of data and material

The data that support the findings of this study are available on request from the corresponding author.

## Data Availability

The data that support the findings of this study are available upon reasonable request to the corresponding author.

